# Readiness, Equity, and Ethical Concerns in Artificial Intelligence (AI) Adoption in Ghana: Implications for AI Integration in Healthcare and Education

**DOI:** 10.64898/2026.02.05.26345694

**Authors:** Gloria Aidoo-Frimpong, Eric Owusu, Dominic Awini Asitanga, Gideon Aduku, Stephen E. Moore, Maame Araba A. Oduro, Zhao Ni

## Abstract

Artificial intelligence (AI) is increasingly positioned as a transformative tool in education and health. Yet empirical evidence on AI readiness in low-and middle-income countries, particularly among youth, remains scarce. This study examined patterns of adoption, equity determinants, and ethical awareness among Ghanaian youth to inform responsible AI integration in education and health systems. A cross-sectional survey was conducted among 200 youth aged 18–35 years in Ghana. Descriptive statistics, chi-square tests, and logistic-regression analyses were used to assess AI adoption, equity patterns, and predictors of readiness. Most participants reported current (89%) or prior (65%) use of AI tools. Accessibility was a significant positive predictor of adoption (β = 0.142, *p* = 0.001), whereas limited internet connectivity (β = –0.088, *p* = 0.049) and perceived exclusion or inequity (β = –0.109, *p* = 0.026) were significant negative predictors. Gender and age differences indicated persistent digital inequities. Ethical concerns were widespread: 51% were somewhat concerned and 39% very concerned about data privacy, algorithmic bias, and transparency. Ghanaian youth exhibit high AI readiness, but it is distributed in structurally uneven and ethically contested contexts. Readiness is best understood as a dynamic interaction between technical access, social inclusion, and trust. Translating readiness into equitable implementation will require investments in digital infrastructure, ethical governance, and participatory design. This study provides one of the first quantitative assessments of AI readiness among African youth and offers an evidence base for developing trustworthy, inclusive AI applications, such as healthcare and educational chatbots, that are grounded in local realities.

**Author Summary:** Artificial intelligence (AI) is often presented as a solution to challenges in healthcare and education. However, there remains limited evidence on people’s readiness to use AI in low-and middle-income countries and on the ways in which equity and ethics shape that readiness. We surveyed 200 youth in Ghana to understand their use of AI tools, perceptions of fairness and ethical concerns. Most participants were already using AI, yet adoption was uneven. Access to reliable internet and devices strongly increased use, while perceptions of exclusion and limited connectivity reduced it. Many youths expressed concern about data privacy, bias, and transparency in AI systems. These findings show that Ghanaian youth are eager but cautious adopters who value fairness and accountability. Building equitable and trustworthy AI systems in education and health will require improving access, addressing social inequalities, and involving youth directly in the design and governance of new technologies.

## Introduction

Artificial intelligence (AI) is rapidly reconfiguring the organization of knowledge, communication, and care worldwide. From clinical decision-support systems to large language models capable of real-time translation and behavioral health counseling, AI tools are now shaping how people access, interpret, and act on information (1–3). While the global diffusion of AI continues to accelerate, empirical research on AI remains uneven across geographic and social (4, 5) Most data informing adoption, regulation, and ethical governance are derived from high-income regions or countries, leaving significant gaps in understanding how AI is perceived, accessed, and governed in low-and middle-income countries (LMICs) (5). In LMICs, readiness to engage with AI (referred to as “AI readiness” in this paper) has often been conceptualized in narrow technocratic terms, emphasizing infrastructure development, computational capacity, and technical skills (6). However, recent global and regional frameworks have underscored that AI readiness extends beyond technical capacity to include social, ethical, and institutional conditions. For example, the World Health Organization’s Ethics and Governance of Artificial Intelligence for Health framework and UNESCO’s Recommendation on the Ethics of Artificial Intelligence emphasize trust, inclusion, accountability, and equity as foundational prerequisites for responsible AI deployment (7, 8). Similarly, scholarship on AI governance and ethics highlights that readiness is shaped not only by digital access and awareness, but also by structural factors such as equitable connectivity, policy protections, data governance, and the ethical climate in which technologies are introduced (5, 9). In the absence of attention to these contextual determinants, AI may entrench existing hierarchies of inequality, widening rather than narrowing health and educational disparities worldwide.

In high-income regions, AI has already been integrated into education and health systems in increasingly formalized ways. In education, AI-powered tools are used to support personalized learning, automated assessment, language translation, and academic advising, with growing institutional guidance on responsible use in schools and universities (10–12). In health systems, AI applications range from clinical decision support and diagnostic imaging to digital triage, mental health chatbots, and population-level surveillance tools, often operating within established regulatory and ethical frameworks (1, 2, 7, 13, 14). These deployments are typically accompanied by investments in infrastructure, workforce training, data governance, and ethical oversight, reflecting a maturation of AI ecosystems that extend beyond experimentation to institutional adoption. However, even in these contexts, concerns about bias, transparency, and equity persist, underscoring that technical sophistication alone does not resolve the social and ethical challenges posed by AI (4, 9, 14, 15). However, the conditions under which AI is being adopted in low-and middle-income countries differ markedly from those in high-income regions. In many LMICs, AI tools are entering everyday educational and health-related spaces in the absence of fully developed infrastructure, governance frameworks, or institutional guidance. This creates a distinct context in which population-level engagement with AI often precedes the formalization of ethical oversight, regulatory safeguards, and equity-oriented policies. Within this broader landscape, Ghana offers a strategically important case for examining how AI readiness takes shape under conditions of rapid diffusion and partial institutional preparedness.

Located in sub-Saharan Africa, Ghana has made substantial investments in digital infrastructure, information and communications technology (ICT) education, and innovation policy, positioning itself as one of West Africa’s fastest-growing technological markets (16, 17).

Additionally, initiatives such as the Digital Ghana Agenda and youth-focused ICT programs have accelerated exposure to AI-powered tools across educational and creative sectors (18). However, these advances coexist with persistent inequities in access, infrastructure reliability, and institutional capacity (17). Internet penetration remains uneven across geographic regions, genders, and socioeconomic statuses, and the cost of connectivity continues to shape patterns of digital participation in Ghana. At the policy level, Ghana is still developing comprehensive frameworks for data governance, algorithmic accountability, and AI ethics, reflecting broader continental trends toward experimentation without fully institutionalized oversight (19). This context of high enthusiasm and partial preparedness illuminates a central paradox of digital transformation in LMICs: technological diffusion often precedes the governance and ethical structures necessary to promote equitable use. Ghana, therefore, is an ideal place to study “frontier readiness,” a stage in which populations are engaging with AI in the absence of mature regulatory ecosystems.

Examining how Ghanaian youth navigate AI tools offers insight into how AI readiness, equity, and ethical awareness emerge organically, shaped by lived experience rather than policy design (20, 21).

Youth, broadly defined in Ghana as individuals aged 15–35 years, represent a critical population for understanding the socio-technical foundations of AI readiness in Ghana and comparable low-and middle-income country contexts. While international definitions of youth vary, with the United Nations commonly using ages 15–24, regional frameworks such as those of the African Union and Ghana’s National Youth Authority extend this range to 35 years, reflecting prolonged transitions in education, employment, and digital engagement (22). The present study focuses on youth aged 18–35 years, aligning with national and regional definitions. As one of the most digitally engaged segments of the population, Ghanaian youth are both early adopters and critical evaluators of emerging technologies, shaping adoption patterns while navigating constraints related to access, cost, and trust (16, 23) Their engagement with AI does not occur in isolation but takes place within social, educational, and economic systems that shape what “readiness” means in practice. Focusing on youth, therefore, provides a window into how digital transformation unfolds at the intersection of individual agency and structural opportunity. In public health and global health contexts, youth are also a priority population for digital interventions, including AI-based approaches to health communication, behavioral counseling, and educational support (24).

Understanding their exposure, access, and ethical awareness can help anticipate the facilitators and barriers that shape the responsible implementation of AI-powered tools. Documenting these early adoption trends is particularly valuable in countries like Ghana, where national strategies for digital transformation are advancing more rapidly than population-level evidence on readiness, equity, or ethical concern (25). This descriptive focus positions the study as an essential starting point for future work examining how youth engagement evolves as AI becomes more embedded in education and health systems. By providing empirical data on adoption, equity, and ethical awareness among Ghanaian youth, the study helps build the foundational evidence base needed to inform contextually appropriate, socially responsive approaches to AI integration.

This study seeks to generate foundational evidence on AI adoption and readiness among youth in Ghana, focusing on the intersection of technological access, social equity, and ethical awareness. Specifically, we aimed to:

1. Assess the level of AI adoption and readiness among Ghanaian youth.
2. Examine how equity-related factors, including gender and access, influence adoption.
3. Explore ethical concerns shaping perceptions of AI use.

By documenting these early patterns, the study provides one of the first population-level snapshots of AI engagement in Sub-Saharan Africa. The findings are intended to inform future research and policy aimed at ensuring that AI systems introduced in education and health are inclusive, contextually grounded, and responsive to the populations they are designed to serve.

## Results

### Demographics

A total of 200 youth participated in the study. Among them, the majority, were 75.0% (n = 150), were aged 18–25 years, while 25.0% (n = 50) were 26 years or older, indicating a predominantly young sample. The majority identified as male (73.5%, n = 147), compared with 26.5% (n = 53) who identified as female.

### AI adoption and readiness

Among the 200 youth surveyed, 89.0% (n = 178) reported current use of AI tools, and 65.0% (n = 130) indicated prior use (Table 1). The most frequently used tools were ChatGPT (50.0%), Quillbot (14.6%), and Photoshop (10.1%). A majority (57.5%) stated that AI-related resources such as internet connectivity, training, or support were “somewhat available.” In comparison, 14.0% reported that resources to support AI uptake and integration into society were “fully available.” Most respondents (64.5%) anticipated that youth would respond positively to broader AI integration.

**Table 1:** Adoption and Readiness (N = 200) Adoption & Readiness Response Category N (%)

| Adoption & Readiness | Response Category | N (%) |
| --- | --- | --- |
| *Current Use of AI Tools* | Yes | 178 (89.0) |
|  | No | 22 (11.0) |
| *Previous Use of AI Tools* | Yes | 130 (65.0) |
|  | No | 70 (35.0) |
| *Resource Availability* | Fully Available | 28 (14.0) |
|  | Somewhat Available | 115 (57.5) |
|  | Not Available | 57 (28.5) |
| *Predicted Reaction* | Positive | 129 (64.5) |
|  | Neutral | 63 (31.5) |
|  | Negative | 8 (4.0) |

### Demographic differences in AI adoption

As shown in Table 2, significant demographic differences were observed in patterns of AI use among Ghanaian youth. Younger participants (18–25 years) were more likely to report active AI use than those aged 25 years and older (χ² = 4.87, *p* = 0.03). This indicates that younger youth are early adopters, likely reflecting greater familiarity with emerging digital platforms and integration of technology into daily activities. Gender differences were also notable; Male participants reported substantially higher AI use (73.5%) compared with females (26.5%), a statistically significant disparity (χ² = 4.54, *p* = 0.05).

**Table 2.**
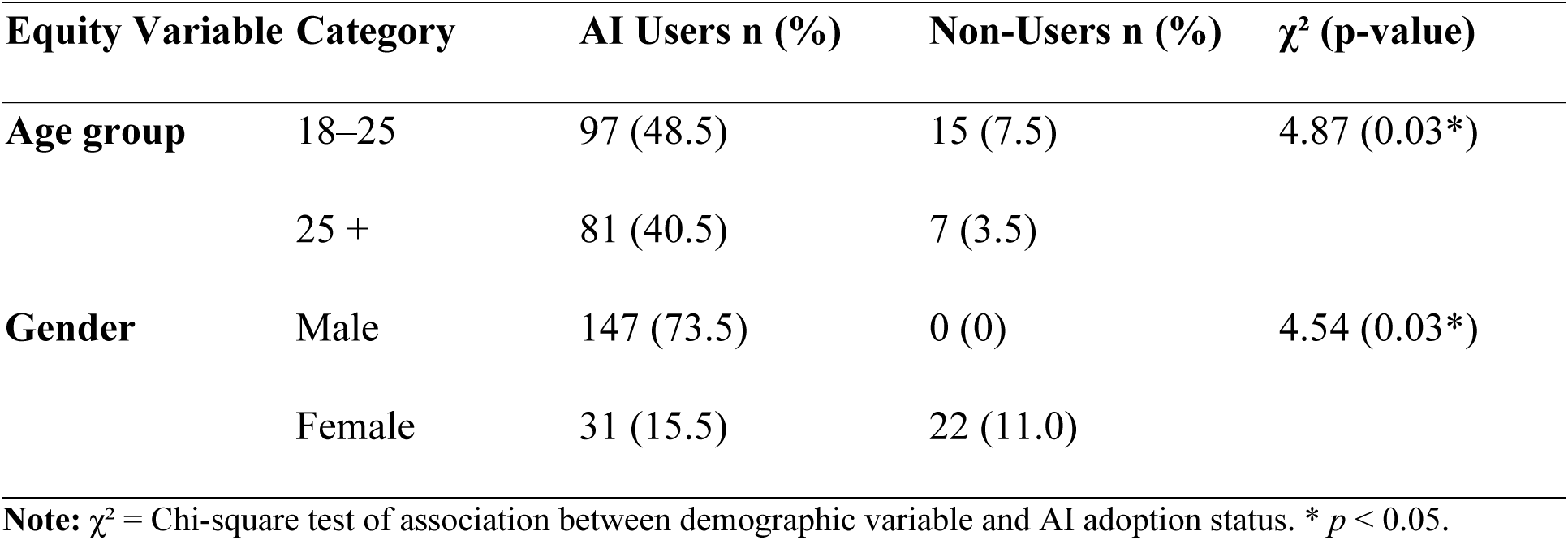
Demographic equity patterns in AI adoption among Ghanaian youth (N = 200)

### Perceived barriers related to equity in AI adoption

Youth identified multiple structural and contextual barriers that could shape equitable AI use (Table 3). Limited internet connectivity was salient, with nearly three-quarters (73.5%) rating it as at least “challenging.” Cost of implementation was also widely viewed as a barrier, with 67.5% describing it as challenging or very challenging. Institutional and governance-related factors were perceived as important. Seventy percent of respondents rated government policies and support as a challenge, and 62.5% identified gaps in data protection and privacy laws as challenging or very challenging. Skills and expertise, education and training, and digital literacy were also frequently described as barriers, although at somewhat lower levels compared with connectivity and cost.

**Table 3:** Perceived Barriers Related to Equity in AI Adoption Among Ghanaian Youth (N = 200)

| Equity-Related Barrier | Not Challenging at All n (%) | Challenging n (%) | Very Challenging n (%) | Extremely Challenging n (%) | Mean ± SD |
| --- | --- | --- | --- | --- | --- |
| Limited Internet Connectivity | 53 (26.5) | 99 (49.5) | 48 (24.0) | 0 (0.0) | 26.5 ± 1.5 |
| Accessibility | 117 (58.5) | 70 (35.0) | 13 (6.5) | 0 (0.0) | 25.3 ± 1.6 |
| Data Centers / Cloud Services | 76 (38.0) | 86 (43.0) | 38 (19.0) | 0 (0.0) | 26.1 ± 1.7 |
| Cost of Implementation | 65 (32.5) | 89 (44.5) | 46 (23.0) | 0 (0.0) | 26.3 ± 1.6 |
| Skills and Expertise | 118 (59.0) | 75 (37.5) | 7 (3.5) | 0 (0.0) | 25.3 ± 1.5 |
| Government Policies and Support | 60 (30.0) | 110 (55.0) | 30 (15.0) | 0 (0.0) | 26.3 ± 1.5 |
| Data Protection and Privacy Laws | 75 (37.5) | 102 (51.0) | 23 (11.5) | 0 (0.0) | 26.0 ± 1.6 |
| Education and Training | 121 (60.5) | 56 (28.0) | 23 (11.5) | 0 (0.0) | 25.3 ± 1.6 |
| Digital Literacy | 107 (53.5) | 66 (33.0) | 27 (13.5) | 0 (0.0) | 25.5 ± 1.7 |
| Cultural Sensitivity | 89 (44.5) | 98 (49.0) | 13 (6.5) | 0 (0.0) | 25.7 ± 1.6 |
| Inclusion and Equity | 102 (51.0) | 90 (45.0) | 8 (4.0) | 0 (0.0) | 25.5 ± 1.6 |

Perceptions related to fairness and inclusion, including cultural sensitivity and “inclusion and equity,” were identified as meaningful but somewhat less intense barriers, with about half of respondents rating these domains as challenging.

### Ethical concerns regarding AI adoption among Ghanaian youth

Ethical concerns were widespread among Ghanaian youth (Table 4). More than half (51.0%) reported being *somewhat concerned* and 39.0% *very concerned* about AI use (*p* < 0.0001). The most frequently cited areas of concern were data protection and privacy (algorithmic bias and fairness), and transparenc**y** of AI decision-making processes. For instance, 51.0% rated privacy issues as a major concern, and nearly half expressed strong concerns about bias and transparency. These findings reveal that, despite high enthusiasm for AI adoption, youth remain alert to its ethical risks. Their emphasis on privacy, fairness, and transparency points to an emerging ethical awareness that should guide the development of governance frameworks for responsible AI integration in education and health.

**Table 4.** Ethical concerns regarding AI adoption among Ghanaian youth (N = 200)

| Area of Ethical Concern | Not Concerned | Somewhat Concerned | Very Concerned | Mean (± |
| --- | --- | --- | --- | --- |
|  | n (%) | n (%) | n (%) | SD) |
| Data protection & privacy | 75 (37.5) | 102 (51.0) | 23 (11.5) | 26.0 ± 1.6 |
| Cultural sensitivity | 89 (44.5) | 98 (49.0) | 13 (6.5) | 25.7 ± 1.6 |
| Transparency of AI systems | 76 (38.0) | 86 (43.0) | 38 (19.0) | 26.1 ± 1.7 |
| Overall level of ethical concern | 20 (10.0) | 102 (51.0) | 78 (39.0) | — |

### Predictors of AI Adoption

Finally, we examined associations between perceived structural and contextual barriers and current AI use using a multivariable logistic regression model (Table 5). Three factors were significantly associated with AI use. Limited internet connectivity was associated with lower odds of current AI use (β = −0.088, OR = 0.92, p = 0.049), and perceived challenges related to inclusion and equity were also associated with reduced odds of AI use (β = −0.109, OR = 0.90, p = 0.026). In contrast, higher perceived accessibility was associated with increased odds of AI use (β = 0.142, OR = 1.15, p = 0.001). Other predictors, including cost of implementation, skills and expertise, government policies and support, data protection and privacy laws, education and training, digital literacy, and cultural sensitivity, were not statistically significant in this model. Results were robust to alternative model specifications, including treating age as a categorical variable and using alternative operationalizations of AI use; no substantive changes in direction or statistical significance were observed.

**Table 5:** Predictors of AI Use among Ghanaian Youth (N = 200)

| Predictor Variable | (β) | (SE) | (OR) | p-value |
| --- | --- | --- | --- | --- |
| Limited Internet Connectivity | −0.088 | 0.044 | 0.92 | 0.049* |
| Accessibility | 0.142 | 0.044 | 1.15 | 0.001* |
| Inclusion & Equity | −0.109 | 0.049 | 0.90 | 0.026* |
| Data Centers & Cloud Services | −0.012 | 0.044 | 0.99 | 0.777 |
| Cost of Implementation | 0.031 | 0.047 | 1.03 | 0.515 |
| Skills & Expertise | 0.067 | 0.041 | 1.07 | 0.104 |
| Government Policies & Support | 0.058 | 0.040 | 1.06 | 0.146 |
| Data Protection & Privacy Laws | 0.060 | 0.043 | 1.06 | 0.158 |
| Education & Training | 0.070 | 0.047 | 1.07 | 0.138 |
| Digital Literacy | −0.059 | 0.043 | 0.94 | 0.168 |
| Cultural Sensitivity | −0.015 | 0.042 | 0.99 | 0.725 |

<sup>a</sup> OR = exp(β), \* p < 0.05.

## Discussion

### Primary Findings

This study contributes new empirical evidence on artificial intelligence readiness among youth in Ghana, a critical but understudied population in sub-Saharan Africa’s digital transformation. Rather than viewing readiness as a purely technical phenomenon, the findings illustrate that it emerges from an interplay of access, inclusion, and ethical trust. Young people demonstrate substantial interest in AI-powered tools, but their engagement is shaped by the structural and relational conditions under which AI becomes available to them. This perspective advances the growing body of work that conceptualizes AI readiness in low-and middle-income settings as a multidimensional capability rather than a fixed skillset.

A central contribution of this study is the identification of readiness as a structural construct.

Our findings suggest that AI readiness among Ghanaian youth is both promising and partial, rooted in high digital enthusiasm but constrained by structural limitations. These findings are consistent with recent studies showing that Ghana youth and students are digitally enthusiastic but face significant structural barriers to sustain engagement with AI and digital technologies (20, 26, 27). The concept of readiness, often framed as a matter of skills or awareness, is better understood here as a contextual capability shaped by material conditions, institutional commitments, and opportunities for meaningful use. The widespread but uneven availability of AI-related resources (somewhat available” for most participants) reflects what scholars describe as *latent readiness*: potential that exists but remains underrealized due to infrastructural and economic constraints (5, 28) Accessibility’s strong positive association with adoption highlights that readiness is not simply about exposure to AI but about the conditions that enable continuity of engagement, stable internet connections, affordable data, and functional devices. This distinction is particularly critical for low-and middle-income countries, where connectivity costs, unreliable electricity, and device sharing can fragment participation (28, 29). In this context, interventions that assume readiness as an individual attribute risk deepening inequity; efforts to strengthen infrastructure, reduce connectivity costs, and expand protected access points are essential for enabling equitable participation in AI ecosystems.

The analysis also highlights important demographic and relational inequities. Differences in adoption by gender and age mirror well-documented digital divides in Ghana and other African contexts. These disparities reflect the social and economic gradients that shape opportunities to engage with technology and suggest that AI innovation does not diffuse evenly across youth populations. Male and younger participants reported higher adoption, consistent with prior research showing that structural determinants, economic resources, social norms, and digital capital shape the opportunity to engage with technology (30, 31). These disparities are not merely demographic artifacts but indicators of unequal capacity to benefit from technological change. The significant negative association between perceived exclusion and AI use adds a relational dimension to this pattern. Youth who viewed AI opportunities as inequitable were less likely to adopt, suggesting that perceptions of fairness and belonging are integral to digital participation (29, 32). In other words, equitable access involves both tangible infrastructure and a sense of social inclusion. This insight aligns with equity frameworks in global health and digital development, which argue that technology adoption is a social process mediated by trust, representation, and recognition (28, 33). Addressing these inequities will require more than generic digital-skills initiatives. Programs that explicitly target underrepresented youth, especially young women and those outside major urban centers, should integrate capacity-building with participatory design, mentorship, and locally relevant content. Policies that reduce data costs, expand public internet access, and incentivize inclusive AI education can help correct the structural asymmetries observed here. Without such interventions, the expansion of AI in Ghana risks reproducing, rather than redressing, existing social hierarchies.

Ethical awareness emerged as another defining feature of youth engagement with AI. Ethical awareness among Ghanaian youth was strikingly high, positioning them not only as early adopters of AI but as critical evaluators of its social and moral implications. Participants expressed sustained concern about three interconnected domains: data privacy, algorithmic bias, and transparency, which mirror global debates on responsible AI. Yet their articulation of these issues situates ethics not in abstract principles but in lived experiences of inequality, surveillance, and exclusion (34, 35). For these youth, ethical concern signals a demand for accountable technology: systems that can be trusted to protect personal data, explain their decisions, and operate fairly across social groups.

These findings challenge assumptions that users in low-and middle-income settings are primarily constrained by access or literacy. Instead, it suggests that ethical literacy coexists with infrastructural limitations. Youth are aware of the risks of digital expansion without adequate governance and view ethics as integral to trust, not peripheral to it (34). Such awareness is crucial as AI applications, especially chatbots and learning platforms, become more embedded in health and education. Responsibility, in this context, extends beyond data protection laws. It requires an inclusive governance model that centers youth voices in defining what “responsible AI” means in Ghanaian society. Youth perspectives offer clear guidance: AI systems in education and health must incorporate transparent consent processes, protections against algorithmic bias, and culturally relevant accountability mechanisms. Embedding these mechanisms in the design and deployment of AI systems will ensure that ethical readiness evolves alongside technical capacity. Rather than framing ethical concern as a barrier to adoption, it should be recognized as evidence of civic maturity and an opportunity to co-produce trustworthy AI ecosystems.

### Implications for AI in Education and Health Systems

The study findings demonstrate that Ghana’s youth are not only active users of emerging technologies but also key stakeholders in shaping the trajectory of AI integration across social sectors. Their readiness to engage with AI, tempered by concerns about access, equity, and ethics, has significant implications for both education and health systems. These sectors are increasingly turning to AI for solutions that promise efficiency, personalization, and reach. Nevertheless, as the findings underscore, readiness is contingent on systemic preparedness, including digital infrastructure, governance, and participatory design.

In education, the widespread familiarity with AI tools such as ChatGPT and Quillbot suggests that youth are already informally integrating AI into their learning processes. This organic adoption presents an opportunity for educational institutions to formalize and scaffold AI use within curricula rather than treating it as peripheral or prohibited (11, 36) AI systems can enhance personalized learning (10), automate feedback (37), and support language translation, capabilities, particularly valuable in multilingual classrooms (36). However, without institutional guidelines, training, and equity safeguards, these tools may deepen existing disparities between students with reliable internet access and those without. Educational policy needs to focus on AI literacy and governance, ensuring that both students and educators understand the capabilities and limitations of AI technologies. Embedding critical digital literacy into teacher training, expanding public access to computing resources, and developing contextually grounded AI ethics curricula would enable youth to use AI creatively and responsibly. These steps would transform AI from an informal learning aid into a structured, equitable educational resource.

The health implications are equally significant. Youth represent both direct beneficiaries and influential intermediaries in the diffusion of AI-enabled health innovations (38). As Ghana and other African countries invest in digital health systems, integrating AI responsibly could improve disease surveillance, health communication, and service delivery. For instance, predictive analytics and natural-language processing can enhance early detection of public health threats, while AI-driven triage tools can help make the best use of limited clinical resources. However, these benefits depend on trust, explainability, and inclusion, areas where our participants’ ethical concerns resonate most strongly. The study’s findings on privacy, fairness, and transparency suggest that young people are already thinking critically about the conditions under which they would share personal information with digital systems.

In the health domain, this awareness translates into concrete design imperatives: ensuring that AI tools collect only necessary data, provide clear consent mechanisms, and allow users to understand and question algorithmic outputs. Local adaptation is also crucial. Algorithms trained on Western data or biomedical assumptions may not reflect Ghana’s epidemiological, linguistic, or cultural context, leading to errors or exclusion. Developing local data ecosystems and ethical review mechanisms will therefore be essential for building trustworthy, high-performing AI systems in health care. In this context, AI chatbots are a useful and scalable way to put these ideas into action. Chatbots can bridge gaps in access to information and care, particularly in areas such as sexual and reproductive health, HIV prevention, mental health, and health-seeking behaviors (39, 40). When designed ethically and inclusively (41), chatbots can democratize access to credible information while preserving privacy through anonymous interaction (42). However, if implemented without safeguards, they risk misinformation, bias, or breaches of confidentiality, concerns that mirror those voiced by participants in this study. The readiness and ethical awareness documented among Ghanaian youth position them as ideal co-design partners in developing such systems. Involving youth in defining chatbot functionality, tone, and privacy standards can ensure that AI applications reflect their realities and values (43). This participatory approach can also serve as a blueprint for broader AI initiatives in education and health, anchoring innovation in local knowledge, transparency, and shared accountability.

### Strengths, Limitations, and Directions for Future Research

This study contributes some of the earliest quantitative evidence on AI readiness, equity, and ethical awareness among youth in Ghana, offering insights with both academic and policy relevance. Its strength lies in its focus on a demographically critical and digitally active population, its use of empirical data from a low-and middle-income country, and its integration of equity and ethics within the broader framework of readiness. By capturing how structural and perceptual factors, such as accessibility, inclusion, and ethical concern, intersect to shape adoption, the study advances a more holistic and context-sensitive understanding of AI readiness than previous literature centered primarily on technical capacity or user attitudes. However, several limitations should be noted. The cross-sectional design precludes causal inference, and the sample reflects youth with some level of digital access, potentially underrepresenting marginalized or rural populations with limited connectivity. Self-reported measures may also introduce recall or social desirability bias, particularly in estimating frequency and comfort with AI use. Furthermore, while the study identifies relationships among readiness, equity, and ethics, it cannot fully explain the mechanisms through which these factors interact over time or across diverse social groups. Future studies using mixed-methods and longitudinal designs could unpack these pathways more precisely, examining, for instance, how improvements in infrastructure or shifts in trust influence patterns of adoption. Future research should also include out-of-school, rural, and informal-sector populations, whose experiences of digital readiness and ethical concern may differ substantially. Comparative research across African contexts could illuminate regional patterns and policy variations affecting AI adoption. Qualitative studies could explore how youth conceptualize fairness, privacy, and accountability in everyday interactions with AI, providing a deeper understanding of ethical readiness as a social process rather than a fixed attitude. Finally, there is a need for implementation-focused research that tests specific interventions derived from these findings, such as participatory co-design of AI chatbots for health communication, inclusive AI literacy programs in schools, or infrastructure-sharing models that reduce connectivity costs.

Evaluating these strategies in real-world settings will help translate empirical readiness into actionable pathways for responsible AI integration across education, health, and governance systems.

## Contribution to literature

This study extends the global evidence base on artificial intelligence in low-and middle-income settings by providing one of the first empirically grounded analyses of AI readiness through an equity and ethics lens. Whereas prior work has often conceptualized readiness as a technical or behavioral attribute, this research reframes it as a multidimensional construct shaped by structural access, perceptions of inclusion, and ethical consciousness. By demonstrating that accessibility and perceived fairness significantly predict adoption, the study provides quantitative evidence that social context, not only infrastructure, determines readiness for AI integration. Additionally, the study’s emphasis on Ghanaian youth introduces essential geographic and demographic diversity to a body of literature predominantly comprised of research from high-income nations and adult professionals. It highlights African youth as both early adopters and ethical actors, capable of evaluating the fairness and risks of AI systems. The inclusion of ethical awareness as a measurable component of readiness offers a conceptual bridge between digital equity research and the emerging field of AI for social benefit.

## Methods

### Study design

This is a cross-sectional survey conducted among 200 Ghanaian youths. Data were collected in community and youth-serving settings across Ghana between May and August 2024. Surveys were administered in locations with variable digital access, which informed the dual online and paper administration.

### Participants

Eligible participants were Ghanaian youth aged 18–35 years, consistent with Ghana’s national definition of youth, which follows the African Union framework defining youth as individuals aged 15–35 years. Participants were required to demonstrate basic English proficiency, defined as the ability to read and understand items written at approximately a primary school (Class 3) level. This requirement was necessary because the questionnaire was administered in English. Exclusion criteria included individuals outside the specified age range, non-Ghanaian residents, and individuals unable to provide informed consent. A total of 200 youth completed the survey.

Recruitment was conducted in Accra and Kumasi, Ghana’s two largest metropolitan areas. These cities serve as major economic, educational, and technological hubs and attract youth from diverse geographic, socioeconomic, and cultural backgrounds across the country. As such, they offer a relevant metropolitan context for examining AI adoption, equity, and ethical perceptions among youth in settings with high digital exposure. A hybrid sampling approach was employed, combining purposive recruitment in educational and community-based settings with random selection of individuals within those settings to support variation by age and gender. Participants provided informed consent prior to participation.

### Data collection

The team developed a structured questionnaire to capture demographics, AI adoption, perceived benefits and challenges, readiness and accessibility, and ethical concerns. Surveys were administered online via Qualtrics, where connectivity allowed, or by paper, where digital access was limited. Paper surveys were double-entered and cross-checked prior to analysis. All data were de-identified before analysis.

### Measures

*Sociodemographic characteristics:* Participants reported their age and gender. Age was collected as a continuous variable and categorized for descriptive analyses. Gender was self-reported.

*AI adoption:* The primary outcome variable was AI adoption, measured by self-reported use of AI-based tools. Current AI use was assessed with the item: *“Are you currently using any AI-based tools?”* (yes/no). Current use was defined as active use of at least one AI-powered tool at the time of survey completion. Prior AI use was assessed using the item: *“Have you ever used any AI-based tools in the past?”* (yes/no), capturing experience with AI tools before the survey period, regardless of current use status. Participants who reported current or prior AI use were asked to identify the specific AI tools they had used.

*Resource availability:* Perceived availability of resources to support AI use was measured using a single-item question assessing access to enabling conditions such as internet connectivity, training, and technical support. Response options were *not available*, *somewhat available*, and *fully available*.

*Anticipated response to AI integration:* Participants’ perceptions of how youth would respond to increased AI integration in education and society were measured using a categorical item with response options *positive*, *neutral*, or *negative*.

*Willingness to adopt AI:* Willingness to continue adopting or supporting AI technologies was measured using a dichotomous item (*willing* vs. *neutral/not willing*), reflecting participants’ self-reported openness to future AI engagement.

*Perceived barriers related to equity in AI adoption*: Participants assessed a series of potential structural and contextual barriers to equitable AI adoption, including internet connectivity, accessibility, cost of implementation, skills and expertise, education and training, digital literacy, government policies and support, data protection and privacy laws, cultural sensitivity, and inclusion and equity. Each barrier was rated on a four-category ordinal scale: *not challenging at all*, *challenging*, *very challenging*, or *extremely challenging*.

*Ethical concerns regarding AI use:* Ethical concerns related to AI technologies were assessed using a single-item measure asking participants how concerned they were about ethical issues associated with AI use. Response options included *not concerned*, *somewhat concerned*, and *very concerned*. Additional items captured specific domains of ethical concern, including data protection and privacy, transparency of AI systems, and cultural sensitivity.

### Data management and statistical analysis

Data were inspected for completeness and internal consistency before locking the analytic file. For descriptive analyses, we summarized categorical variables with frequencies and percentages and continuous variables with means and standard deviations. The primary outcome, AI use, was coded as a binary indicator of active AI tool use (yes/no). To examine factors associated with AI use, we fit a multivariable logistic regression model including prespecified predictors reflecting structural and equity-related barriers, including accessibility, internet connectivity, and perceived inclusion and equity. Predictor variables were selected a priori based on conceptual relevance to AI use and the study’s research questions, rather than through bivariate screening.. Models adjusted for age and gender. We report odds ratios (ORs) with 95% confidence intervals (CIs); statistical significance was set at p < 0.05. Analyses were conducted in IBM SPSS Statistics version 27 (44). Because this was an exploratory, observational study with no anticipated effect size, no a priori power calculation was performed. Sensitivity analyses were performed by refitting models with age specified as both a continuous and categorical variable and by assessing alternative operationalizations of AI adoption where applicable, to evaluate the robustness of findings.

## Ethical considerations

The study protocol was reviewed and approved by the SUNY Brockport Research Ethics Committee (Protocol No. STUDY00004950). In addition, local ethical approval was obtained from the Kwame Nkrumah University of Science and Technology Institutional Review Board (CABE/CD/DF/32) in Ghana. All participants provided written informed consent. The de-identified dataset used for analysis contained no direct personal identifiers.

## Conclusion

This study shows that Ghanaian youth are not only active users of AI-powered tools but critical stakeholders in shaping the conditions under which AI can be integrated into education and health. Their readiness is promising yet contingent, emerging where digital access, social inclusion, and ethical trust intersect. Accessibility remains the strongest facilitator of adoption, while perceptions of inequity and concerns about privacy, bias, and transparency shape how youth evaluate and engage with AI systems. These patterns illustrate that digital transformation cannot be driven by technology alone; readiness is a social and ethical achievement grounded in the environments that make sustained and equitable engagement possible. Efforts to expand AI in Ghana must be paired with investments in infrastructure, governance, and participatory design. Reducing structural barriers, addressing gender and age disparities, strengthening data-protection frameworks, and embedding fairness in algorithmic design are essential for building trust and ensuring equitable benefit. Youth participation should be viewed not as optional but as foundational to the legitimacy and effectiveness of AI-enabled systems.

## Data Availability

The de-identified dataset generated and analyzed during the current study is available from the corresponding author upon reasonable request and with appropriate institutional approvals, due to ethical restrictions related to participant privacy.

